# Midbrain structure volume, estimated myelin and functional connectivity in idiopathic generalised epilepsy

**DOI:** 10.1101/2022.08.23.22278902

**Authors:** Andrea McKavanagh, Adam Ridzuan-Allen, Barbara A.K. Kreilkamp, Yachin Chen, José V. Manjón, Pierrick Coupé, Martyn Bracewell, Kumar Das, Peter N. Taylor, Anthony G. Marson, Simon S. Keller

## Abstract

**Background:** Previous neuroimaging studies reported functional and structural impairments of the upper basal ganglia (specifically, the striatum) in idiopathic generalised epilepsy (IGE). However, these studies often overlook lower basal ganglia structures located in and adjacent to the midbrain due to poor contrast on clinically acquired volumetric T1-weighted scans. Here, we acquired 3D isotropic T1-weighted, T2-weighted and resting state functional MR images to investigate differences in volume, estimated myelin content and functional connectivity in three midbrain and midbrain-adjacent structures in patients with IGE: the substantia nigra (SN), subthalamic nuclei (SubTN) and red nuclei (RN).

**Methods:** A total of 33 patients with IGE (23 refractory, 10 non-refractory) and 39 age and sex matched healthy controls underwent MR imaging. The SN, SubTN and RN were automatically segmented from T2-weighted images. Estimated median myelin content for each structure was determined using a previously described T1-weighted/T2-weighted ratio method. Functional connectivity analysis between midbrain structures and the rest of the brain was performed using seed-based resting state fMRI analysis and compared between participant groups using the CONN toolbox running in SPM12 (pFDR<0.05, cluster corrected).

**Results:** An increased volume of the right RN was found in patients with IGE relative to healthy controls. Structural volumes of the right SubTN differed between patients with non-refractory and refractory IGE. No myelin alterations of midbrain structures were found in patients with IGE or between treatment outcome groups. Functional connectivity alterations were found for all midbrain regions in patients with IGE, including significantly decreased functional connectivity between the left SN and the thalamus compared to controls, and significantly increased functional connectivity observed between the right SubTN and the left superior frontal gyrus in patients with IGE relative to controls. Connectivity alterations specific to patients with non-refractory IGE were also found.

**Conclusion:** We report volumetric and functional connectivity alterations of midbrain and lower basal ganglia structures in patients with IGE. We postulate that an increased structural volume of the RN in patients is due to increased iron deposition that impacts on T2-weighted contrast. These findings are consistent with previous experiential work demonstrating pathophysiological abnormalities of the lower basal ganglia in animal models of generalised epilepsy.

## Introduction

Idiopathic generalised epilepsy (IGE) is a group of generalised epilepsy sub-syndromes which accounts for one third of all epilepsies. The disorders are characterised by a presumed genetic predisposition, generalised spike/poly-spike wave discharges on EEG and no identifiable brain lesion on routine MRI (Scheffer *et al*., 2017). IGEs are network disorders characterised by aberrant structural and functional brain networks which are thought to promote generalised ictal activity (Liao *et al*., 2013; Caeyenberghs *et al*., 2015; Liu *et al*., 2017; Larivière *et al*., 2020).

The basal ganglia (BG) are a cluster of interconnected subcortical nuclei that have been reported to play prominent roles in IGE ictal networks (Luo *et al*., 2012; Dong *et al*., 2016; Gong *et al*., 2021). The BG act as critical hubs, receiving, relaying, and modulating information to and from the cortex via various neural pathways which include motor, sensory, dorsolateral prefrontal, lateral orbitofrontal and anterior cingulate pathways (Graybiel *et al*., 1994; Nicola, 2007; Seger and Spiering, 2011).

Previous animal studies showed BG involvement in the propagation of epileptic discharges (Deransart *et al*., 1998, 2000; Slaght *et al*., 2004). Furthermore, neuroimaging studies have reported morphometric (Seeck *et al*., 2005; Du *et al*., 2011; Keller *et al*., 2011; Luo *et al*., 2011; Saini *et al*., 2013; Kim *et al*., 2018; Zhong *et al*., 2018), diffusion (Keller *et al*., 2011; Luo *et al*., 2011; Yang *et al*., 2012) and functional (Luo *et al*., 2012; Rektor *et al*., 2013; Gong *et al*., 2021; Zhang *et al*., 2021) MRI alterations of the BG in patients with IGE relative to healthy controls. Despite the clear involvement of BG nuclei in IGE, the focus of studies has almost entirely been on upper BG structures, such as the striatum and globus pallidus, to the exclusion of lower BG structures in the midbrain.

Lower BG structures include the substantia nigra (SN) and the subthalamic nucleus (SubTN). The primary reason that morphometric analyses of these structures in IGE are lacking is due to the poor contrast of these structures on clinically acquired isotropic T1-weighted images. The full anatomical profile of these structures, along with other midbrain structures such as the red nucleus (RN), are preferentially observed with isotropic T2-weighted images, which are not routinely acquired in patients with IGE. Despite this, there is convincing experimental evidence for the involvement of these structures in IGE.

Increased agonistic activation of nigral GABA receptors inhibits induced generalised motor seizure activity in male Sprague-Dawley rats (Iadarola and Gale, 1982). Conversely, convulsive activity was not reduced when GABA receptor activation was increased in the striatum, hippocampus, or thalamus, suggesting that the SN is important for the inhibition of generalised seizures (Iadarola and Gale, 1982). Other work reported that a reduction of inhibitory GABAergic neurons in BG structures including the SN pars reticula (SNr) is likely to contribute to increased seizure susceptibility of flurothyl-induced seizures in a genetic mouse model (*Brd +/-)* of JME (Velíšek *et al*., 2011), further highlighting how the SN may be implicated in genetic models of generalised epilepsy. Additional to modifications of the GABAergic system in the lower midbrain, alterations to the dopaminergic system have been reported in patients with IGE. Reduced dopamine transporter (DAT) binding in the midbrain and specifically in the SN was found in patients with JME using Positron Emission Tomography (Ciumas *et al*., 2008). However, no changes were observed in striatal DAT binding (Ciumas *et al*., 2008). Other computational work has reported that the SN and SubTN are part of an integrated network that modulates the EEG presentation of absence seizures (Hu *et al*., 2020). Furthermore, despite few studies linking the RN with generalised epilepsy, some animal work has suggested that the RN may have a role in the expression of generalised seizures. For example, GTCSs were triggered in a convulsant mouse model by low threshold stimulation of the RN and high threshold stimulation of SN (Gioanni, Gioanni and Mitrovic, 1991). Other work suggested that the RN may have a role in seizure inhibition during induced tonic-clonic seizures (Fernández-Guardiola *et al*., 1974). There is therefore evidence to suggest that midbrain structures could play an important role in the pathophysiology of IGE.

In the present study we prospectively acquired MRI data using a multi-sequence protocol that included 3D isotropic T1-weighted (T1W), T2-weighted (T2W) and resting-state functional MRI (rsfMRI). Data were acquired from patients with refractory (continued seizures despite anti-seizure medication) and non-refractory (seizure free) IGE patients and healthy controls. We used these data to investigate whether patients with IGE showed evidence of altered volume, estimated myelin and functional connectivity of SN, SubTN and RN. Furthermore, we explored whether SN, SubTN and RN alterations are potential markers of pharmacoresistence by comparing imaging data between patients with refractory and non-refractory IGE.

## Methods

### Participants

Thirty-three patients with IGE and thirty-nine age and sex matched healthy controls with no history of neurological or psychiatric disease were recruited into this study. Demographic information can be found in Table 1. All patients were diagnosed with IGE by an experienced neurologist based on the ILAE classification criteria (Scheffer *et al*., 2017), including patient history, seizure semiology, and characteristic generalised spike/polyspike wave discharges on EEG. Ten patients were categorised as non-refractory, defined as being seizure free for 12 months at the time of recruitment (Kwan *et al*., 2010). Refractory patients experienced two or more seizures during the 12 months immediately prior to recruitment (Kwan *et al*., 2010) and had failed more than two anti-seizure medication regimes. For further details on patient cohort see supplementary material (Table S1). Participants were scanned at the Department of Neuroradiology at The Walton Centre NHS Foundation Trust and informed written consent was obtained for each (local research ethical committee reference 14/NW/0332).

**Table 1.** Participant demographic and clinical characteristics

| Groups |  |  |  |  | Stats |  |
| --- | --- | --- | --- | --- | --- | --- |
|  | *Healthy Controls* | *All Patients* | *Ref* | *Non-Ref* | HC vs All | Ref vs Non-Ref |
| N | 39 | 33 | 23 | 10 | - | - |
| Sex F/M (n, %) | 23(59)/16(41) | 18(55)/15(45) | 13(56)/10(44) | 5(50)/5(50) | 0.81 | >0.99 |
| Mean age (±SD) | 32(8.6) | 32(15) | 35(15) | 26(11) | 0.13 | 0.08 |
| Mean age at Onset (±SD) | - | 13(5.6) | 14(5.4) | 12(5.9) | - | 0.61 |
| Mean epilepsy Duration (±SD) | - | 19(16) | 21(16) | 14(16) | - | 0.10 |

Sex significance calculated using Fisher's Exact test and Age/Onset/Duration significance using Mann Whitney U test; Ref = Refractory; Non-ref = Non-refractory; Age/Onset and Duration in years; significance = $p < 0.05$

### Image Acquisition

Each participant underwent an MRI protocol that consisted of T1W, T2W and rsfMRI sequences using a 3T GE Discovery MR 750 MRI scanner with a 32-channel head coil: (1) T1W; fast spoiled gradient echo (FSPGR) images with Phased Array Uniformity Enhancement (PURE) signal inhomogeneity correction (140 slices, TR=8.2 ms, TI=450 ms, TE=3.22 ms, flip angle=12, with 1mm isotropic voxel size, acquisition time: 3:48 mins); (2) T2W; T2W CUBE images (with PURE correction, 312 slices, TR=2500 ms, TI = N/A, TE=71.2ms, flip angle=90, with 0.5 mm isotropic voxel size); (3) rsfMRI; T2W Pulse sequence gradient echo, TE = 25 ms; TR = 2,000 ms; FOV= 24; slice thickness = 2.4 mm; voxel size = 3.75mm x 3.75mm; 180 volumes; 38 slices; flip angle = 75, acquisition time: 6 mins. All participants were awake and instructed to look at a visual fixation point during scanning acquisition.

### Segmentation and Volumetry

Figure 1 provides an overview of the image analysis pipeline used in the present study. Automatic segmentation software pBrain was used to automatically segment the SN, SubTN and RN from T2W scans (Manjón *et al*., 2020) through the online volBrain platform (Manjón and Coupé, 2016). The software efficiently and accurately segments structures using multi-atlas label fusion technology on T2W scans as deep brain structures are more easily detectable on T2W scans than other imaging modalities (Manjón *et al*., 2020). Prior to segmentation, T2W images were rigidly aligned to T1W images using FSL (FMRIB Software Library v6.0; https://fsl.fmrib.ox.ac.uk/fsl/fslwiki) FLIRT (Jenkinson *et al*., 2002). Co-registered T2W images were then processed using pBrain and volumes of associated structures were calculated by the segmentation tool.

**Figure 1.**
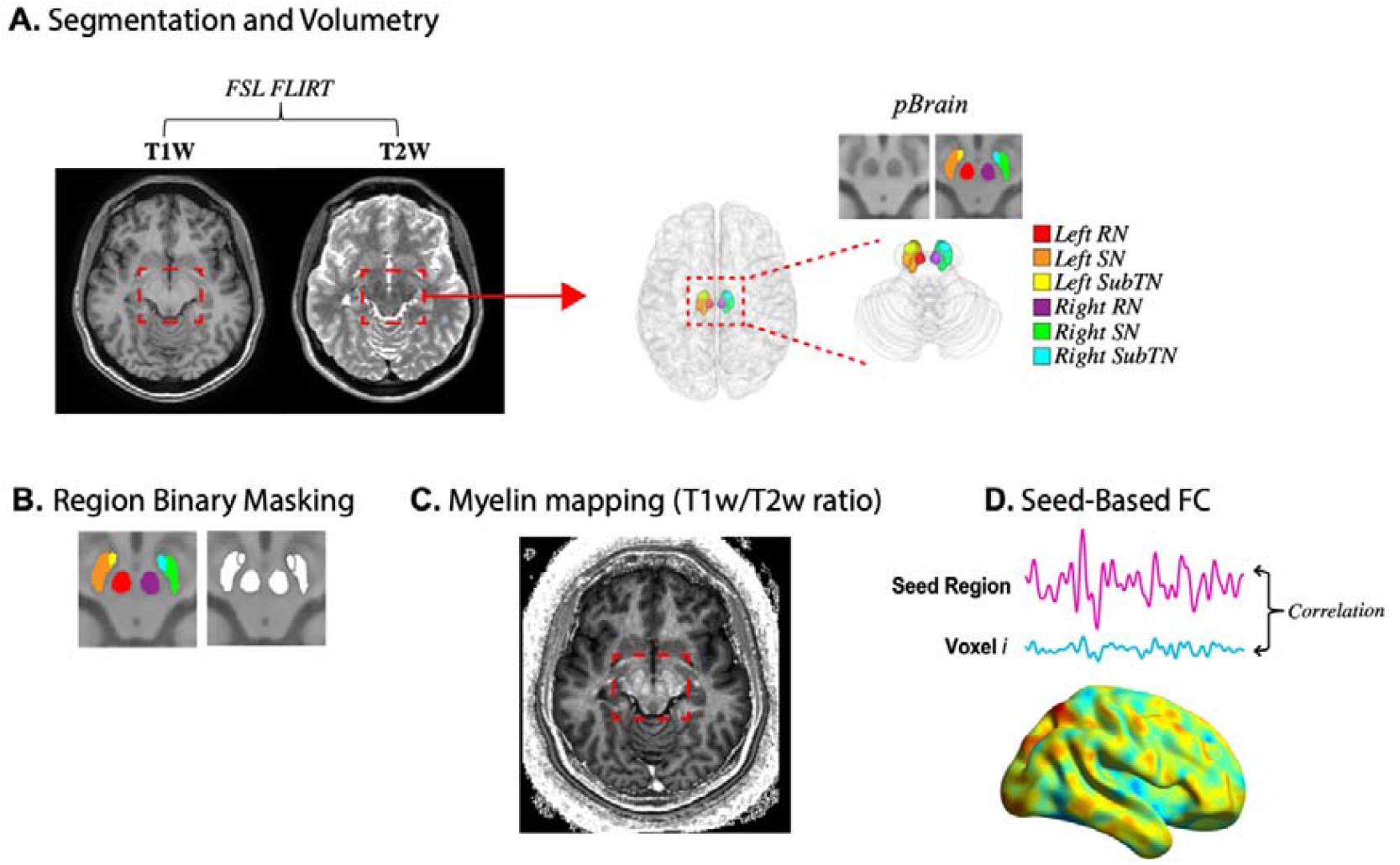
Methods pipeline schematic. (A) T2W images are rigidly aligned to T1W images prior to input into automatic segmentation software pBrain, which segments and computes midbrain structure volumes. (B) Segmented regions are thresholded, binarized and masked. (C) Myelin estimation maps are generated from a T1W/T2W ratio computation. Regional masks are overlayed onto myelin maps to extract overall myelin content per region per subject using FSL utilities. (D) Structural masks are used as seeds in rsfMRI seed-based functional connectivity analysis.

### Constructing binary masks

Binary masks of segmentations were generated using FSL utilities to be used for further downstream myelin and connectivity analysis (Figure 1.B). Structures were assigned a numerical label from the segmentation software which was used as an absolute threshold to remove voxels with intensity values above/below the thresholded value. Subsequently, thresholded segments were binarized using FSL utilities by retaining non-zero voxels only, generating binary masks for all structures. All binary masks were checked manually for segmentation accuracy.

### Estimated myelin content

Myelin content was estimated using a whole brain myelin mapping technique (Glasser and Van Essen, 2011). MRI contrast in cortical grey matter is influenced by iron depositions, and myelin content and iron are colocalised (Fukunaga *et al*., 2010). It has therefore been suggested that grey matter myelin content can be indirectly measured using MRI signal intensities (van der Weijden *et al*., 2021). Grey matter myelin inversely covaries with T1W and T2W signal intensity, whilst background noise does not correlate between the two modalities. Thus, Glasser and colleagues proposed a straightforward non-invasive methodology for measuring estimated myelin content using a ratio of T1W/T2W image intensities (Equation 1), whilst removing the need for bias field correction due to inhomogeneous MRI signals, and enhancing the contrast to noise ratio for myelin content (Glasser and Van Essen, 2011):

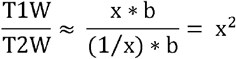

Equation 1. Where x = myelin content in T1W image, 1/ x = myelin content in T2W image (inverse of T1W), b = receive bias field in both images, and x^2^ = enhanced myelin contrast image (T1W/T2W image).

Image pre-processing and whole brain myelin mapping were carried out according to the previously described method (Glasser and Van Essen, 2011). Subsequently, lower basal ganglia and midbrain binary masks were applied to the myelin enhanced T1W/T2W images (Figure 1.C) and the median voxel signal values within each mask were calculated using FSL utilities to determine the myelin content per segmented structure.

### Statistical analysis of volumetry and myelin estimation

Permutation testing via FSL toolbox PALM (Winkler *et al*., 2014) was used to compare midbrain volume/myelin content between groups. The software constructs a standard linear model for the observed parameters and calculates a t-statistic for each contrast (IGE vs controls; non-refractory vs refractory), then the model is permuted (10,000 iterations) and a p value computed (FDR corrected). All parameters were corrected for age and sex using a GLM prior to permutation testing, and volumes were also corrected for ICV (as calculated by pBrain).

### Resting State fMRI pre-processing

rsfMRI data were pre-processed using Statistical Parametric Mapping software (SPM12, Welcome Trust Centre for Neuroimaging, London, UK; https://www.fil.ion.ucl.ac.uk/spm/). Pre-processing steps included slice time correction of the 180 volumes using the first volume as a reference. Head motion and EPI distortion were corrected using the spatial realignment and unwarp tools, respectively. A motion artefact threshold (translation > 3 mm, rotation > 1^°^) was applied which caused the exclusion of five controls due to excessive head motion. Subsequently, scans were spatially normalised to the ICBM 152 template using an affine registration and then smoothed with an 8mm full width at half maxima (FWHM) gaussian kernel to increase signal to noise ratio. T1W images were skull stripped and segmented into grey matter, white matter, and cerebrospinal fluid (CSF) using the SPM Computational Anatomy Toolbox (CAT12; http://www.neuro.uni□jena.de/cat/) and all segmentations were non-linearly registered to the ICBM 152 template. Pre-processing was completed using the SPM Functional Connectivity Toolbox (CONN; https://web.conn-toolbox.org, (Whitfield-Gabrieli and Nieto-Castanon, 2012) integrated in SPM12. As part of CONN’s default denoising pipeline to prevent potential confounding effects, nuisance variables including the first 5 principal components of white matter and CSF signals, and 6 motion parameters including their associated first order derivates, were linearly regressed out using Ordinary Least Squares (OLS) regression. Furthermore, a band-pass filter (0.01-0.08 Hz) was applied to control for scanner drift and high-frequency physiological noise (Smith *et al*., 1999; Cordes *et al*., 2001; Murphy, Birn and Bandettini, 2013).

### Seed-based functional connectivity and statistical analysis

Seed-based connectivity (SBC) maps for each participant were computed between midbrain seeds and all other grey matter voxels. First, masks were linearly transformed to MNI space using SPM normalise and registration accuracy was checked manually. Standard space masks were then imported into the CONN toolbox and used as seeds of interest in the rsfMRI seed-based functional connectivity. A Fisher-transformed correlation coefficient between the average BOLD signal of the seed and the BOLD signal of every grey mater voxel within the rest of the brain was calculated. Cluster level statistics using Random Field Theory (Worsley *et al*., 1996) were applied in CONN to the SBC maps of each midbrain region to compare results between subject groups (IGE vs controls; non-refractory vs refractory). A statistical parametric map (corrected for age and sex) of t-values estimated using a GLM was generated and thresholded to p<0.001 to define clusters. The size of each cluster (number of voxels) was measured and compared to a known probability density function to give each cluster a p-value which underwent multiple comparison correction using the false discovery rate (FDR). Significance for clusters was thresholded at p<0.05 (FDR-corrected).

## Results

### Volumetry and estimated myelin findings

Patients with IGE had an increased volume of the right RN (t = 2.11, p = 0.03, pFDR = 0.15) compared to controls (Figure 2.A; Table 2).

**Table 2.** Comparison of volumes between subject group pairs using FSL PALM.

|  | All Patients<br>(N = 33) |  | Refractory<br>(N = 23) |  | Non-Refractory<br>(N = 10) |  | Healthy Controls<br>(N = 39) |  | All Patients vs<br>Controls |  |  | Non-Refractory<br>vs Refractory |  |  |
| --- | --- | --- | --- | --- | --- | --- | --- | --- | --- | --- | --- | --- | --- | --- |
| *Region* | *Mean volume*<br>*(cm<sup>3</sup>)* | *SD* | *Mean*<br>*volume*<br>*(cm<sup>3</sup>)* | *SD* | *Mean*<br>*volume*<br>*(cm<sup>3</sup>)* | *SD* | *Mean*<br>*volume*<br>*(cm<sup>3</sup>)* | *SD* | *tval* | *pval* | *pFDR* | *tval* | *pval* | *pFDR* |
| Left SN | 0.54 | 0.15 | 0.53 | 0.17 | 0.54 | 0.10 | 0.50 | 0.08 | 1.09 | 0.28 | 0.42 | -0.32 | 0.75 | 0.99 |
| Right SN | 0.49 | 0.11 | 0.48 | 0.12 | 0.51 | 0.10 | 0.44 | 0.08 | 1.88 | 0.06 | 0.15 | 0.02 | 0.99 | 0.99 |
| Left SubTN | 0.11 | 0.07 | 0.11 | 0.08 | 0.12 | 0.02 | 0.09 | 0.02 | 1.00 | 0.36 | 0.43 | -0.90 | 0.32 | 0.96 |
| Right SubTN | 0.11 | 0.03 | 0.11 | 0.04 | 0.11 | 0.02 | 0.10 | 0.02 | -0.06 | 0.95 | 0.95 | -2.12 | *0.04 | 0.23 |
| Left RN | 0.26 | 0.06 | 0.25 | 0.06 | 0.29 | 0.06 | 0.24 | 0.04 | 1.80 | 0.07 | 0.15 | 0.34 | 0.73 | 0.99 |
| Right RN | 0.25 | 0.07 | 0.25 | 0.07 | 0.27 | 0.07 | 0.23 | 0.04 | 2.11 | *0.03 | 0.15 | 0.14 | 0.89 | 0.99 |

*SN* = Substantia Nigra; *SubTN* = Subthalamic Nucleus; *RN* = Red Nucleus; *SD* = standard deviation; *FDR* = False discovery rate; Significance = $p < 0.05$ ; \* = uncorrected significance.

**Figure 2.**
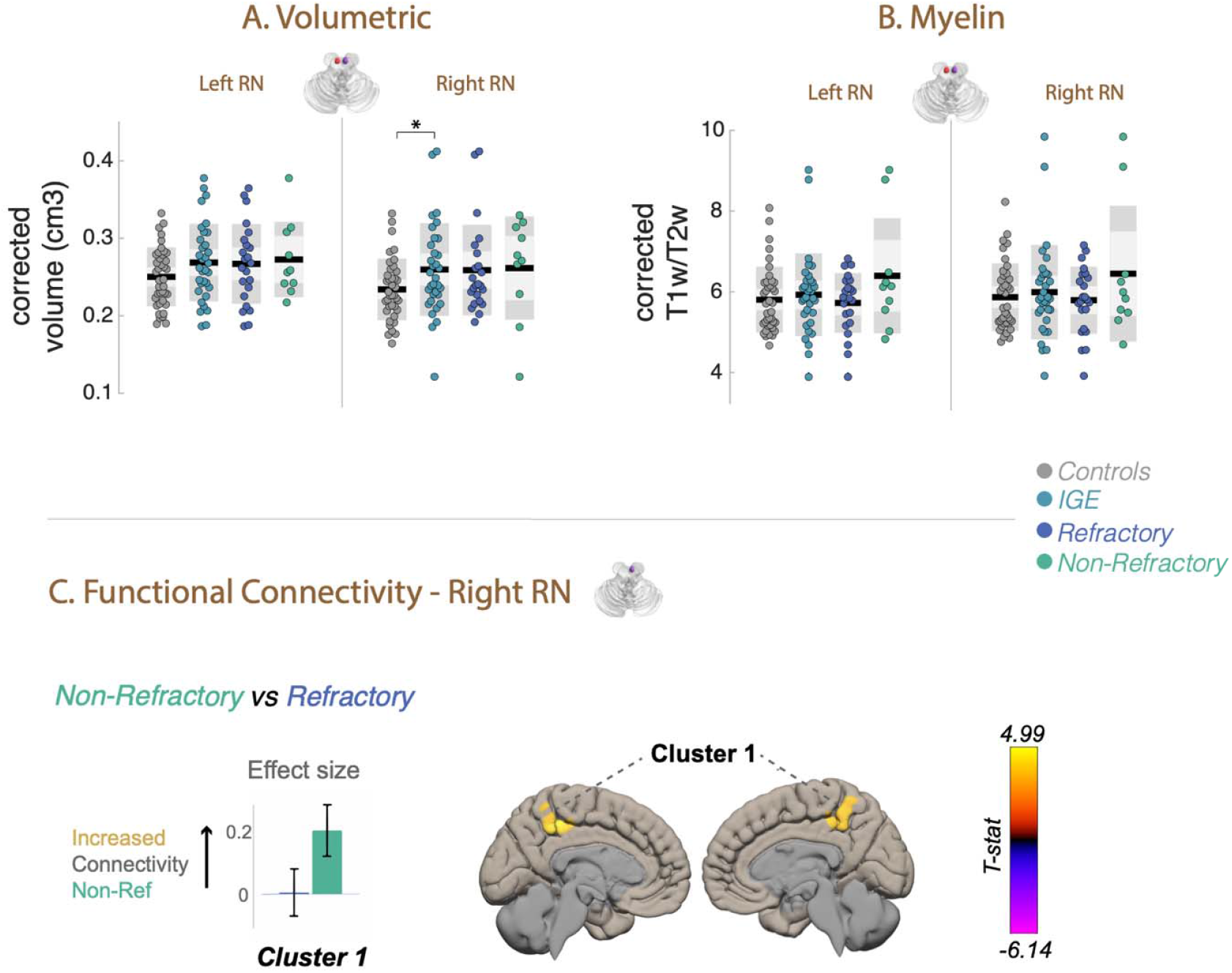
Volume, myelin estimation and functional connectivity of the red nucleus (RN) bilaterally in patients with IGE. (A) Volumes (cm^3^) of the left and right RN for subject groups. All volumes were corrected for age, sex and ICV. (B) Myelin estimation (ratio T1W/T2W) of the left and right RN for subject groups. Estimated myelin was corrected for age and sex. (C) Functional connectivity differences (pFDR<0.05, cluster corrected) of the right RN between patients with non-refractory and refractory IGE. Bar charts show effect size differences, 90% confidence intervals, and direction of contrast for subject group pairs. Cluster colour indicates t-score and trend direction (increased/decreased connectivity). (RN= red nucleus; *=p<0.05)

Further, patients with non-refractory IGE had a decreased volume of the right SubTN (t = - 2.12, p = 0.04, pFDR = 0.23) relative to patients with non-refractory IGE (Figure 3.A; Table 2).

**Figure 3.**
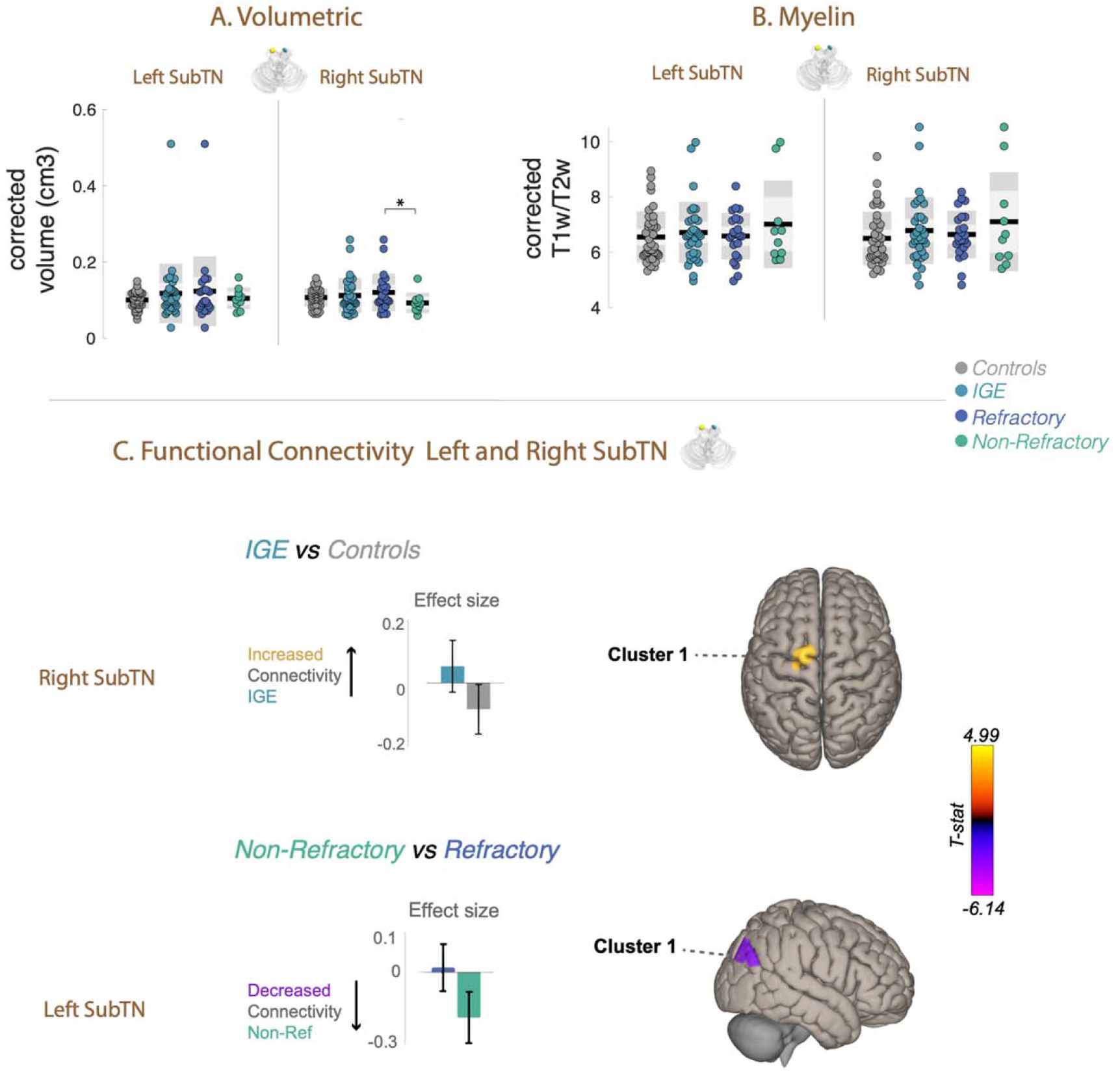
Volume, myelin estimation and functional connectivity of the subthalamic nucleus (SubTN) bilaterally in patients with IGE. (A) Volumes (cm^3^) of the left and right SubTN for subject groups. All volumes were corrected for age, sex and ICV. (B) Myelin estimation (ratio T1W/T2W) of the left and right SubTN for subject groups. Estimated myelin was corrected for age and sex. (C) Functional connectivity differences (pFDR<0.05, cluster corrected) of the right SubTN of patients with IGE relative to controls, and of the left SubTN between patients with non-refractory and refractory IGE. Bar charts show effect size differences, 90% confidence intervals, and direction of contrast for subject group pairs. Cluster colour indicates t-score and trend direction (increased/decreased connectivity). (SubTN = Subthalamic Nucleus).

No volume alterations of the SN and SubTN were found between patients with IGE and controls, and no alterations of the SN and RN were found between patients with refractory and non-refractory IGE (Table 2). Furthermore, no significant myelin alterations of the midbrain (Figure 2.B, Figure 3.B, and Figure 4.B; Table 3) were found in patients with IGE relative to controls or between patients with refractory and non-refractory IGE

**Table 3.** Comparisons of myelin content between subject group pairs using FSL PALM.

|  | All Patients<br>(N = 33) |  | Refractory<br>(N = 23) |  | Non-Refractory<br>(N = 10) |  | Healthy Controls<br>(N = 39) |  | All Patients vs<br>Controls |  |  | Non-Refractory<br>vs Refractory |  |  |
| --- | --- | --- | --- | --- | --- | --- | --- | --- | --- | --- | --- | --- | --- | --- |
| *Region* | *Mean*<br>*T1W/T2*<br>*W* | *SD* | *Mean*<br>*T1W/T2W* | *SD* | *Mean*<br>*T1W/T2W* | *SD* | *Mean*<br>*T1W/T2W* | *SD* | *tval* | *pval* | *pFDR* | *tval* | *pval* | *pFDR* |
| Left SN | 5.93 | 1.16 | 5.90 | 0.96 | 6.01 | 1.60 | 5.70 | 0.90 | 1.22 | 0.23 | 0.33 | 0.91 | 0.36 | 0.36 |
| Right SN | 5.81 | 1.15 | 5.73 | 0.88 | 5.99 | 1.67 | 5.64 | 0.92 | 1.04 | 0.30 | 0.33 | 1.24 | 0.22 | 0.31 |
| Left SubTN | 6.69 | 1.18 | 6.63 | 0.92 | 6.84 | 1.68 | 6.53 | 0.94 | 0.99 | 0.33 | 0.33 | 1.13 | 0.26 | 0.31 |
| Right SubTN | 6.76 | 1.28 | 6.69 | 0.95 | 6.93 | 1.89 | 6.48 | 0.98 | 1.39 | 0.17 | 0.33 | 1.13 | 0.26 | 0.31 |
| Left RN | 5.91 | 1.00 | 5.77 | 0.70 | 6.23 | 1.48 | 5.79 | 0.84 | 1.15 | 0.26 | 0.33 | 1.99 | 0.05 | 0.23 |
| Right RN | 5.98 | 1.14 | 5.84 | 0.78 | 6.28 | 1.74 | 5.85 | 0.84 | 1.04 | 0.31 | 0.33 | 1.76 | 0.08 | 0.23 |

*SN* = Substantia Nigra; *SubTN* = Subthalamic Nucleus; *RN* = Red Nucleus; *SD* = standard deviation; *FDR* = False discovery rate; Significance = $p < 0.0$ ; \* = uncorrected significance.

**Figure 4.**
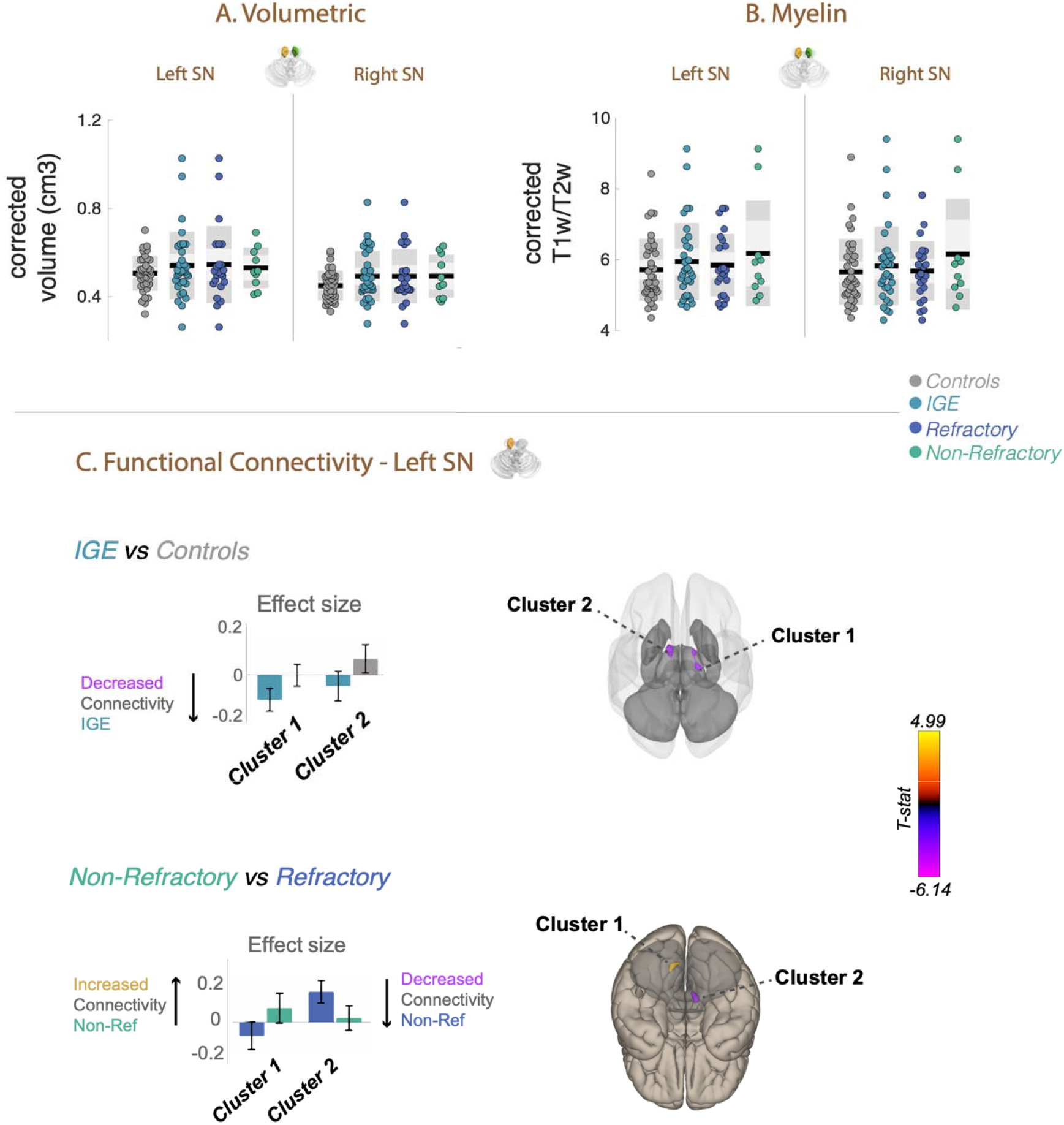
Volume, myelin estimation and functional connectivity of the substantia nigra (SN) bilaterally in patients with IGE. (A) Volumes (cm^3^) of the left and right SN for subject groups. All volumes were corrected for age, sex and ICV. (B) Myelin estimation (ratio T1W/T2W) of the left and right SN for subject groups. Estimated myelin was corrected for age and sex. (C) Functional connectivity differences (pFDR<0.05, cluster corrected) of the right SN between patients with non-refractory and refractory IGE. Bar charts show effect size differences, 90% confidence intervals, and direction of contrast for subject group pairs. Cluster colour indicates t-score and trend direction (increased/decreased connectivity). (SN= substantia nigra, *=p<0.05)

### Group differences in functional connectivity

In patients with IGE, connectivity alterations were found for all midbrain regions (Table 4) and results differed depending on structure laterality (left or right) and the group contrast analysed (IGE compared to controls; non-refractory compared to refractory). Patients with IGE had significantly decreased functional connectivity between the left SN and the bilateral thalamus and caudate nucleus compared to controls (Figure 4.C). Compared to patients with refractory IGE, patients with non-refractory IGE had significantly increased connectivity between the left SN and the left cerebellum, and decreased connectivity between the left SN and the right cerebellum. Significantly increased functional connectivity was observed between the right SubTN and the left superior frontal gyrus in patients with IGE relative to controls (Figure 3.C). Additionally, patients with non-refractory IGE had significantly decreased connectivity between the left SubTN and right lateral occipital cortex compared to patients with refractory IGE. Significantly increased functional connectivity was observed between the right RN and the left and right precuneus cortex in patients with refractory IGE relative to those with non-refractory IGE (Figure 2.C).

**Table 4.**
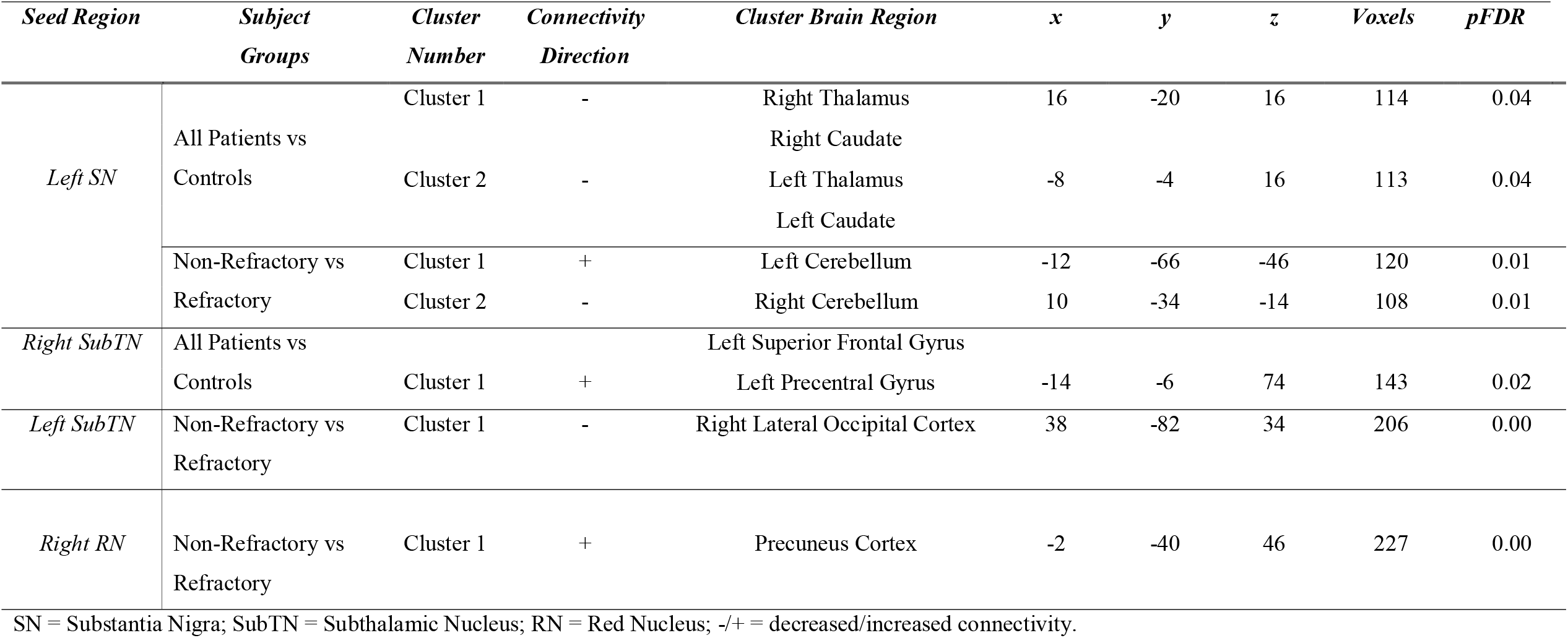
Midbrain Functional Connectivity Results

## Discussion

We investigated whether volume, estimated myelin and functional connectivity of the lower BG structures located within and adjacent to the midbrain are altered in patients with IGE. We observed an increased volume of the RN and functional connectivity alterations of the SN, SubTN and RN in patients with IGE compared to healthy controls. Furthermore, we investigated if differences existed between patients with refractory and non-refractory IGE. We report volumetric alterations between treatment outcome groups in the SubTN and differential patterns of functional connectivity alterations between subcortical structures and other grey matter regions. The clinical and biological implications of these findings are presented prior to discussion of pertinent methodological issues.

Previous studies reported MRI-determined volumetric changes in patients with IGE, which included structures located in the upper BG including the striatum and pallidum (Betting *et al*., 2006; Ciumas and Savic, 2006; Kim *et al*., 2007; Du *et al*., 2011). To our knowledge, we are the first to investigate the structure of the SN, SubTN and RN in patients with IGE using MRI. We found volumetric increases of the RN and of the SubTN in refractory IGE compared to non-refractory IGE which are known to modulate motor movement and play a role in motor disorders (Florio *et al*., 2018; Basile *et al*., 2021), and therefore have the potential to play an important motoric role in the initiation and termination of seizures in epilepsy. Increased regional volumes on T1W images have been commonly found previously in IGE, including in the thalamic nuclei (*c*.*f*. more studies report thalamic volume reduction rather than volume increase, Chen et al., 2021), the right medial frontal gyrus, right anterior cingulate cortex (Bin *et al*., 2017), precentral gyrus and paracentral lobule (Hsin *et al*., 2017). An increase in midline frontoparietal grey matter was reported in early voxel-based studies of IGE (Woermann et al., 1999, 2000). The underlying causes of increased structural volume in IGE are not clear. It has been hypothesised that T1W-derived grey matter increases may have been caused by consequences of functional alterations, or by histopathological alterations such as microdygenesis (Meencke and Janz, 1985; Meencke and Veith, 1999). Despite those hypotheses, there have been no MRI volumetric studies on the SN, SubTN and RN in patients with IGE. Volumetric increases of the RN has been reported previously in Parkinson’s disease and authors have hypothesised volume alterations may be related to an increase of iron content in the region (Camlidag *et al*., 2014). In terms of epilepsy, a review of the literature reported correlations between epilepsy and iron metabolism (Chen *et al*., 2020). An accumulation of iron, altered iron metabolism and iron induced inflammation has been reported in post-mortem brain tissue of patients with TLE (Zimmer *et al*., 2021) and injections of iron into the cortex can cause epileptic seizures in animal models (Willmore *et al*., 1978; Willmore, Sypert and Munson, 1978). The exact pathophysiological correlates of volumetric increases in IGE remain unknown therefore further research of the structures of the lower BG and midbrain nuclei, with the use of high-resolution T2W images, is warranted.

We found no evidence of changes to estimated myelin within midbrain nuclei in patients with IGE. Previous studies reported reduced estimated myelin in patients with epilepsy including right hemisphere epilepsy (Goldsberry *et al*., 2011), febrile epilepsy (Moldovan *et al*., 2018) and refractory epilepsy (Deodato *et al*., 2002). Decreased myelin in the white matter in TLE (Bartolini *et al*., 2017) has been found and decreased myelin water fraction in the frontal lobe white matter in patients with CAE (Drenthen *et al*., 2019). Furthermore, those with disorders of myelin content have been found to have an increased susceptibility to having epileptic seizures (Lapato *et al*., 2017; Gruntz *et al*., 2018; Baker *et al*., 2019; Schorner and Weissert, 2019). Most studies focused on white matter tracts given the extensive axonal myelination in this area of the brain. Diffusion tensor imaging (DTI) studies have frequently been performed in patients with IGE and suggested a potential relationship between altered diffusion properties of white matter tracts and abnormal white matter myelination (Liu *et al*., 2011; Focke *et al*., 2014; Qiu *et al*., 2016; Knake *et al*., 2017). There is evidence to suggest that diffusion scalar metrics are sensitive to myelin content (Chang *et al*., 2017; Zhou *et al*., 2020; Lazari and Lipp, 2021). In contrast to previous white matter DTI studies, we analysed estimated myelin within midbrain grey matter nuclei, which has not been previously performed in patients with IGE. We used a well-established T1W/T2W ratio method which is sensitive to grey matter myelin content (Glasser and Van Essen, 2011; Iwatani *et al*., 2015; Nakamura *et al*., 2017; Luo *et al*., 2019; Tullo *et al*., 2019; Paquola *et al*., 2020). The absence of estimated myelin content differences in the SN, SubTN and RN may be explained by different myelination patterns and myelination functions between white and grey matter (Timmler and Simons, 2019). It has been hypothesised grey matter myelinated axons may be used for metabolic reasons or as inhibitors of axonal sprouting rather than saltatory conduction as seen in the white matter (Timmler and Simons, 2019). Alternatively, it is also possible myelin is not impacted in the grey matter nuclei of the midbrain and lower BG in IGE.

We report decreases in functional connectivity between the SN and the thalamus and caudate in patients with IGE. Diverging results have shown the SN, SubTN, thalamus and caudate as part of the BG resting state network are more intrinsically functionally connected in patients with IGE compared to controls (Luo *et al*., 2012). Alternative results may be explained by different methodological approaches used between studies. Authors Luo et al., use a spatial ICA approach which provides an intrinsic connectivity measure between all structures within a network, whereas we used a localised seed-based temporal correlation analysis which provides a single connectivity measure between specific structures within the network.

Functional alterations found between the midbrain, thalamus and caudate are unsurprising as the SN modulates the direct and indirect pathway to initiate/inhibit motor movement looping signals to the cortex via interactions with the BG (Calabresi *et al*., 2014). Decreased functional connectivity of the SN highlights the weaker integration of these nuclei within motor pathways, which may be a factor in the clinical manifestations of seizures in IGE.

We found increased functional connectivity between the right SubTN and the left superior frontal gyrus in patients with IGE. Probabilistic tractography studies have established projection fibres exist between both structures as part of the hyper direct pathway for the modulation of motor movement (Aravamuthan *et al*., 2007; Lambert *et al*., 2012). The importance of these interconnected white matter tracts for movement has been evidenced by their role in the deep brain stimulation of the SubTN in Parkinson’s disease (Vanegas-Arroyave *et al*., 2016). The superior frontal gyrus has been found to be impaired in patients with IGE, with previously reported volumetric (Deng *et al*., 2019; Kazis *et al*., 2021) and microstructural alterations (Focke *et al*., 2014), and epileptiform discharges (Jun *et al*., 2019). Therefore, abnormal connectivity between the SubTN and superior frontal gyrus may be related to altered networks contributing to the motor manifestations of seizures in IGE.

We also found connectivity alterations characteristic of patients with non-refractory IGE. We observed decreased connectivity between the left SubTN and the right lateral occipital cortex and increased functional connectivity between the right RN and precuneus compared to patients with refractory IGE. Cognitive dysfunction, ongoing cognitive decline and more severe cognitive impairment is found in patients with refractory epilepsy (Jokeit and Ebner, 1999; Vingerhoets, 2006; Li *et al*., 2020). Functional circuits for visuospatial processing involve both the SubTN and occipital lobe in humans (Schmalbach *et al*., 2014). The RN is similarly involved in cognitive circuits including episodic memory function and is functionally connected to the precuneus cortex (Nioche, Cabanis and Habas, 2009; Zhang *et al*., 2015). Furthermore, we observed connectivity alterations between the SN and cerebellum in patients with non-refractory IGE. Various studies found alterations between cerebellum connectivity and BG structures in IGE (Moeller *et al*., 2011; Li *et al*., 2017; Jiang *et al*., 2018). The cerebellum has been reported to be a modulator of generalised seizures (Gotman *et al*., 2005; Kros *et al*., 2015) and motor control projections are found between the dentate nuclei of the cerebellum and the SNr in the midbrain (Milardi *et al*., 2016). These network connectivity alterations may be implicated in cognitive impairments and motor control in IGE and thus in non-refractory patients may suggest possible changes in cognitive and motor circuits indicative of seizure freedom.

## Limitations

There are several limitations of this study. Different medication regimes between patients may influence results as some anti-epileptic drugs can decrease cerebral blood flow (Joo *et al*., 2006) which may impact BOLD signal and functional connectivity. However, this effect may be specific to certain resting state networks as limited anti-epileptic drug effects on functional connectivity of the BG network has been found previously in IGE (Luo *et al*., 2012). Furthermore, our patient sample includes several sub-disorders of IGE which may influence results due to possible syndrome specific pathophysiological mechanisms. However, shared similarities between IGE sub-syndromes including, clinical features (Reutens and Berkovic, 1995), genetic influences (De Kovel *et al*., 2010) and some pathophysiological mechanisms (Benuzzi *et al*., 2012; Masterton, Carney and Jackson, 2012; Kim, Kim and Suh, 2019) have previously been found. Therefore, investigating volumetric, myelin and functional similarities between sub-syndromes is important. Lastly, low spatial resolution of fMRI and the small size of midbrain nuclei investigated may decrease the reliability of the BOLD signal in the midbrain area, therefore studies with an increased spatial resolution could account for this limitation in the future.

## Conclusion

We report significant structural and functional connectivity alterations of midbrain structures in patients with IGE. Patients with IGE have an increased T2W derived volume of the RN of the midbrain, which is likely due to increased iron accumulation in this region. These structural changes may relate to functional connectivity alterations that we observed between lower BG / midbrain structures and other cortical and subcortical grey matter areas. Our study demonstrates both structural and functional alterations of these anatomical regions in patients with IGE.

## Data Availability

All data produced in the present study are available upon reasonable request to the authors

## Funding and Acknowledgments

This work was supported by a UK Medical Research Council DiMeN DTP studentship awarded to AM. SSK acknowledges support from the UK Medical Research Council (Grant Number MR/S00355X/1)

---

**Table S1.** Clinical characteristics of patients.

| Patient | Age | Sex | Onset | Duration | Category | FH | PS | Seizures | ASM(mg/day) |
| --- | --- | --- | --- | --- | --- | --- | --- | --- | --- |
| 1 | 21-25 | F | 11-15 | 9 | Ref | N | N | AS, MS | LEV 300, TOP 300, Clob 10 |
| 2 | 16-20 | M | 16-20 | 3 | Ref | Y | Y | GTCS | VPA 1000 |
| 3 | 16-20 | F | 6-10 | 11 | Ref | Y | N | AS, GTCS | LTG 200 |
| 4 | 21-25 | M | 16-20 | 6 | Non-Ref | N | N | MS | LEV1500, VPA 1600 |
| 5 | 61-65 | F | 11-15 | 47 | Ref | Y | N | AS, GTCS | VPA 2500 |
| 6 | 21-25 | M | 11-15 | 9 | Ref | Y | N | AS, MS, GTCS | CBZ1000, LEV 3000, VPA 2500 |
| 7 | 21-25 | F | 11-15 | 6 | Ref | N | N | AS, MS, GTCS | LEV 4000, VPA 2000 |
| 8 | 31-35 | F | 21-25 | 9 | Ref | Y | N | MS, GTCS | LEV 3500, Clob 15 |
| 9 | 36-40 | M | 16-20 | 20 | Ref | Y | N | GTCS | VPA 600, LTG 50 |
| 10 | 66-70 | M | 26-30 | 38 | Ref | N | N | AS, GTCS | VPA 2000, LTG 200, PB 150, Clob 10 |
| 11 | 46-50 | F | 6-10 | 39 | Ref | N | N | AS | VPA 1200, LTG 200, LEV 2500 |
| 12 | 16-20 | M | 6-10 | 12 | Ref | N | N | GTCS | VPA 2000 |
| 13 | 21-25 | F | 11-15 | 11 | Ref | Y | N | MS, GTCS | TOP 100 |
| 14 | 31-35 | M | 6-10 | 29 | Ref | N | N | GTCS | LEV 2000, VPA 2000 |
| 15 | 16-20 | M | 11-15 | 4 | Ref | N | N | AS, GTCS | VPA 1500, ZON 350 |
| 16 | 36-40 | M | 16-20 | 22 | Ref | Y | Y | GTCS | VPA 1000, LTG 75 |
| 17 | 21-25 | F | 16-20 | 8 | Non-Ref | N | N | AS, GTCS | VPA 1000, LTG 200, LEV 4000 |
| 18 | 21-25 | M | 16-20 | 5 | Ref | N | N | AS, MS, GTCS | VPA 2400 |
| 19 | 36-40 | F | 16-20 | 19 | Ref | N | N | GTCS | LEV 1250, TOP 100 |
| 20 | 31-35 | F | 16-20 | 16 | Ref | N | N | GTCS | LEV 2000, LTG 400 |
| 21 | 31-35 | F | 16-20 | 15 | Ref | N | N | AS, MS, GTCS | VPA 1500, LEV 3500 |
| 22 | 21-25 | M | 16-20 | 7 | Non-Ref | N | N | AS, GTCS | VPA 2100, LEV 500 |
| 23 | 16-20 | F | 11-15 | 6 | Non-Ref | Y | N | GTCS | LEV 3000 |
| 24 | 56-60 | F | 11-15 | 43 | Ref | N | N | GTCS | VPA 1000, ZON 400, Clon 1.5 |
| 25 | 16-20 | F | 11-15 | 3 | Non-Ref | N | Y | AS, MS | LEV 2000 |
| 26 | 21-25 | M | 1-5 | 20 | Non-Ref | N | Y | AS, MS | VPA 1400 |
| 27 | 21-25 | M | 11-15 | 11 | Ref | N | N | MS | VPA 1700 |
| 28 | 56-60 | F | 1-5 | 53 | Non-Ref | N | Y | AS | VPA 1500 |
| 29 | 56-60 | F | 6-10 | 50 | Ref | N | N | AS, GTCS | VPA 1200, CBZ 600 |
| 30 | 31-35 | M | 6-10 | 26 | Non-Ref | N | N | AS | VPA 1800 |
| 31 | 16-20 | F | 11-15 | 5 | Non-Ref | N | N | AS, MS | LEV 1000 |
| 32 | 56-60 | F | 6-10 | 50 | Ref | Y | N | GTCS | VPA 2000, LTG 75 |
| 33 | 16-20 | M | 16-20 | 4 | Non-Ref | N | N | AS, GTCS, MS | VPA 1700, ETX 500 |

Onset = age range of onset of epilepsy in years; Duration = Duration of epilepsy in years; F = female; M = male; Ref = refractory; Non-Ref = Non-Refractory; FH = family history; PS = photosensitive; ASM = anti-seizure medication; AS = absence seizures; GTCS = primary generalized tonic-clonic seizures; MS = myoclonic seizures; VPA = Valproic acid; ZON = Zonisamide; LEV = Levetiracetam; TOP = Topiramate; Clob = Clobazam; Clon = Clonazepam; LTG = Lamotrigine; CBZ = Carbamazepine; PB = Phenobarbital; ETX = Ethosuximide; N = no; Y = yes.

